# e-Health Multimodal Interventions for Older Adults with Chronic Non-Cancer Pain: A Scoping Review Protocol

**DOI:** 10.1101/2023.07.27.23293235

**Authors:** Annalisa De Lucia, Alessandro Chiarotto, Ilenia Pasini, Sara Pachera, Lidia Del Piccolo, Cinzia Perlini, Valeria Donisi

## Abstract

**Background:** Chronic non-cancer pain (CNCP) is one of the most prevalent health conditions among the elderly, with a considerable impact on the person’s physical, mental and social functioning. The use of a bio-psycho-social multidisciplinary approach has become widely recommended for more likely effective management of chronic pain. In recent years, the growing development and application of e-Health (or digital health) within pain medicine has been showing encouraging results. However, the application of such technologies in the field of pain management among elderly is yet understudied, particularly in regard to the potential impacts of multimodal therapies (i.e., interventions which integrate a physical and a psychological component) provided via digital devices.

**Objective:** The overall aim of this scoping review is to systematically map the existing literature about the e-Health multimodal interventions designed for older adults with CNCP.

**Methods:** Multiple electronic databases (PubMed, Cochrane CENTRAL, Web of Science, PsycINFO) will be searched for relevant articles to August 2023. The review will adhere to the Joanna Briggs Institute (JBI) methodology and will utilize the Preferred Reporting Items for Systematic Reviews and Meta-Analyses extension for scoping reviews (PRISMA-ScR) reporting guideline and checklist. All eligible studies will be evaluated against the 16-item Quality Assessment Tool (QATSDD). The extracted information will be presented in tabular form along with a narrative summary that is in line with the scoping review’s objective.

## Introduction

One of the most prevalent health conditions among the elderly is chronic pain, which is frequently accompanied by cognitive decline, sleep difficulties, psychological disorders, and functional impairments (Dagnino & Campos, 2022). According to estimates, the prevalence of pain in community-dwelling older adults ranges from 25% to 50%, and it reaches 80% in institutionalized elders, reflecting a current clinical issue (Cravello et al., 2019). As a result of its higher risk of poor outcomes due to multimorbidity and polypharmacy, the old population poses many challenges to the health care professional in terms of treatment (Niknejad et al., 2018). Thus, clinicians and researchers have recently turned their attention to a bio-psycho-social multidisciplinary approach that considers not only and exclusively biological aspects in the pain management, but also psychological and social factors that have seemed to be involved in the experience of pain (Kamper et al., 2015; Schwan et al., 2019; Perlini et al., 2020; Gandolfi et al., 2021). In fact, using a multimodal approach that includes various combinations of both pharmacological and non-pharmacological therapies (such as psychological interventions, physiotherapy, occupational therapy, complementary and alternative medicine) is recommended for a more likely effective management of chronic pain in older adults (Dale & Stacey, 2016; Cheng et al., 2017). Furthermore, accessibility to effective treatment programs for persistent pain frequently appears to be constrained, as a result of both limitations in the healthcare system (such as long waiting lists and a shortage of qualified professionals) and challenges faced by patients due to physical and mobility limitations, as well as the consequent difficulty in accessing the right healthcare networks. In that sense, a breakthrough has been made with the recent development of e-Health (or digital health), which is defined as “*the use of information and communications technology in support of health and health-related fields*” (WHO, 2019). Digital applications and e-Health interventions span a wide spectrum and are constantly developing. For the management of chronic pain, various e-Health solutions have been presented and implemented. Slattery and colleagues (2019) provided examples of web-based interventions where patients are involved by using a wider range of technological tools (such as computers or mobile devices, telephone-supported, interactive voice response technology, virtual reality (VR), video teleconferencing, and mobile phone applications). The medical application of digital tools has also been recently implemented in pain medicine with encouraging results (Lee, 2021). However, the use of such technologies in the field of pain management among elders is still little explored, especially regarding the potential effects of multimodal interventions (i.e., including both a physical and a psychological component) delivered by digital devices. Since this topic has not yet been comprehensively reviewed and considering the complex and heterogeneous nature of the body of knowledge in that field, a scoping review is thought to be the most appropriate methodology to explore and analyze such an emerging area of research.

## Review questions

This scoping review aims to systematically map the existing literature about the e-Health multimodal interventions, involving both a physical and a psychosocial component, designed for older adults with chronic non-cancer pain (CNCP) in order to answer the following research questions:

1. What is the body of evidence (e.g., in terms of number and quality of studies) and gaps in the current literature?
2. What are the main characteristics of the e-Health multimodal interventions involving physical and psychosocial components in older adults with CNCP?
3. What kinds of populations have been included in the available studies, for example, in terms of specific chronic pain conditions, age sub-groups, comorbidities?
4. Which are the main outcomes and promising results of those interventions?

## Inclusion criteria

### Participants/population

Older adults; 65 years of age and older, since this age has been conventionally accepted as the beginning of old age (e.g. Singh & Bajorek, 2014) and population studies around the world show that the over 65 age group have a higher prevalence of chronic pain than the general adult population (Domenichiello & Ramsden, 2019); no restrictions in terms of gender or in terms of other clinical conditions different from chronic non-cancer pain.

### Concept

e-Health-based (all type of e-Health will be accepted) multimodal interventions, involving both a physical approach component (such as therapeutic exercise, functional training, etc.) and a psychosocial approach component (any intervention targeting one or more emotional, cognitive, behavioral or interpersonal aspects) designed for older adults with CNCP and targeting the following main outcomes: pain (e.g. severity), emotional functioning (e.g. emotional distress, anxiety and depression symptoms, catastrophizing, kinesiophobia, perceived self-efficacy), physical functioning (e.g. functional mobility, endurance) and integrated outcomes (e.g., health-related quality of life, wellbeing, general functioning and disability).

### Context

Studies involving the use of e-Health interventions among older adults with CNCP in any care setting (e.g., primary care, outpatient, community, etc.) will be eligible for inclusion.

### Types of study to be included

Both qualitative and quantitative studies will be included. The following types of studies will be excluded: systematic review, narrative review, meta-analysis, bibliometric analysis, letter, case-study, book/book chapter, comment, editorial, congress abstract or symposium, poster presentation, and dissertation.

## Methods

This scoping review will adhere to the Joanna Briggs Institute (JBI) methodology and will utilize the Preferred Reporting Items for Systematic Reviews and Meta-Analyses extension for scoping reviews (PRISMA-ScR) reporting guideline and checklist (Tricco et al., 2018; Peters et al., 2020; Peters et al., 2022).

### Search strategy

The following electronic databases will be searched for relevant studies: PubMed, Cochrane CENTRAL, Web of Science, PsycINFO.

A combination of key terms related to the following four main topics will be used as a search strategy in the above mentioned databases: 1) *e-Health* (e.g., telemedicine, telehealth, m-Health), 2) *Psychosocial and Physical Interventions* (e.g., psychotherapy, psychoeducation, physiotherapy, physical activity) *or Multimodal Intervention* (e.g., multicomponent/multifactorial intervention, mind-body therapy), 3) *Older People* (e.g., elderly, seniors) and 4) *Chronic Pain* (e.g., chronic pain). Only studies published in English and Italian from inception to August 2023 will be considered.

### Data extraction (selection and coding)

All articles yielded will be exported into the Systematic Reviews Web application Rayyan (Ouzzani et al., 2016) and duplicates will be removed. Two reviewers will independently assess titles and abstracts for eligibility. Full texts of the potentially eligible articles will be retrieved and will be assessed against eligibility criteria by two reviewers. Doubts will be discussed, and, where necessary, a third reviewer will be involved. A systematic and in-depth description of the following data will be done: study design; characteristics of the sampled population for age, gender, type of chronic pain, pain duration, main comorbidities, setting; the intervention outcomes; the type of e-Health tools for delivering the intervention, specifying if the intervention is guided or unguided and who are the providers; the type of intervention, its conceptual basis, structure, main components, duration and format; older adults’ experiences and perceptions of e-Health interventions when applicable; the follow-up duration of the study, when applicable; when involved, the type of control group. Two reviewers will extract data from the selected studies using a data collection form in Microsoft Excel.

### Assessment of methodological quality

All eligible studies will be evaluated against the 16-item Quality Assessment Tool (QATSDD) (Sirriyeh et al., 2012). The tool demonstrates good validity and reliability for evaluating the quality of sets of research papers adopting various methodologies (e.g., qualitative and quantitative). It consists of 16 criteria, each of which is graded from 0 (meaning “not at all”) to 3 (meaning “complete”). For qualitative or quantitative research, the highest score is 42; for mixed-method studies, the maximum score is 48. For each included article, the score assigned to each item and the paper’s overall quality score (i.e., resulting from the sum of individual scores for each indicator) will be reported. Furthermore, in addition to the average quality score for all papers, each item’s mean and standard deviation will be calculated to describe the items with higher and lower values. Two independent raters will assess the quality of the included studies using the QATSDD. Any potential disagreement will be discussed, including a third rater to adjudicate.

### Data analysis and presentation

The extracted information will be presented in tabular form along with a narrative summary that is in line with the scoping review’s objective.

## Data Availability

Not applicable

